# p16 Expression and its correlation with the clinical pathological characteristics of cervical cancer patients: a systematic review and meta-Analysis

**DOI:** 10.1101/2025.02.27.25323000

**Authors:** Le Chong, Zuowei Zou, Luhua Xia, Xinhua Wang, Zhanfei Dong, Yanping Zhao, Youxiang Hou

**Affiliations:** Key Laboratory of Oncology of Xinjiang Uyghur Autonomous Region, Department of Ultrasonography, Tumor Hospital Affiliated to Xinjiang Medical University, China; Department of Nuclear Medicine, National Cancer Center/National Clinical Research Center for Cancer/Cancer Hospital, Chinese Academy of Medical Sciences and Peking Union Medical College, China; Key Laboratory of Oncology of Xinjiang Uyghur Autonomous Region, Department of Nuclear Medicine, Tumor Hospital Affiliated to Xinjiang Medical University, China; Department of Radiation Oncology, Tumor Hospital Affiliated to Xinjiang Medical University, China

## Abstract

**Introduction:** Overexpression of p16 has been reported in various human tumors. However, the correlation between p16 overexpression and the clinical pathological characteristics of cervical cancer remains contentious. Therefore, this meta-analysis aims to systematically evaluate the relationship between p16 expression and the clinical pathological features of cervical cancer.

**Methods:** PubMed, Embase, Cochrane Library (Central), Web of Science (SCI Expanded), and Chine databases (CNKI, VIP, Wanfang, and CBM) were searched for relevant articles published between 1 January 2010 and March 01,2024. Case-control studies related to the correlation between p16 expression and cervical cancer were selected to evaluate the correlation between p16 expression and the clinicopathological features of cervical cancer patients. The software Review Man 5.4 was used for statistical and data processing, and heterogeneity tests and sensitivity analyses were performed. Literature quality was evaluated using the Newcastle-Ottawa scale. The software STATA 16.0 was selected for sensitivity analysis of the included studies, and publication bias was tested via Begg’s test.

**Results:** Twenty-three eligible studies with, 1611 cervical cancer patients and 852 normal controls were included. p16 protein expression (cervical cancer patients VS normal controls) OR=74.32, 95%CI: 50.35-109.69, P<0.05; The expression of p16 protein in cervical cancer patients is associated with lymph node metastasis (metastatic lymph nodes vs. non-metastatic lymph nodes) OR=1.79, 95% CI: 1.29-2.47, P<0.05;The expression of p16 in patients with cervical cancer correlated with the degree of differentiation of the tumours (high and medium differentiation vs. low differentiation) (OR= 0.41, 95% CI: 0.30-0.56, P<0.05); the age of disease onset (age≤40 vs. age>40) (OR=0.55, Z=3.00, P=0.003<0.05), the status of choroidal infiltration in cervical cancer patients (yes vs. no) (OR=3.53, 95% CI: 1.25-10.01, P=0.02 <0.05), and tumour FIGO stage (stage I-II vs. stage III-IV) (OR=0.37, 95% CI: 0.18-0.76, P=0.0.007 <0.05), and these differences were statistically significant. p16 expression was not correlated with infiltration depth, tumour pathology type, tumour size, and these differences were not statistically significant.

**Conclusion:** The p16 protein is closely related to the occurrence and development of cervical cancer and can serve as a reference indicator for the early warning and diagnosis of cervical cancer.

## 1. Introduction

Cervical cancer is one of the three major malignant tumours in gynaecology, and the incidence and mortality rate of this cancer ranks fourth among female cancers [1–3]. Cervical cancer poses a serious threat to women’s health, and it is showing a trend towards rejuvenation in recent years [4]. Cervical cancer screening is a global research hotspot, but pathological diagnosis is not sufficient to assess the severity and prognosis of cervical disease. With the development of molecular biology, the search for more objective molecular markers is highly important for the prevention and treatment of cervical cancer. Some specific tumour biomolecule markers improve the specificity of detecting high-grade cervical lesions and cervical cancer, reduce the misdiagnosis rate of abnormal cervical cytology results and provide valuable information for predicting invasion and metastasis and evaluating the prognosis of patients with cervical cancer [5,6].

The p16 gene belongs to the INK4 family and consists of four members, p16INK4a, p15INK4b, p18INK4C, and p19INK4D, which all share common biological properties, i.e., inhibition of cell growth and tumour suppression. Mutations, deletions, and methylation of the p16 gene play important roles in tumorigenesis, progression, and metastasis [7–9]. p16 loses its cancer inhibitory function due to mutation, deletion or methylation, which lead to abnormalities or obstacles in cell cycle regulation. p16 participates in the occurrence, development and metastasis of oral, lung, pancreatic, cervical and many other tumours, P16 may be an effective target for the diagnosis and treatment of clinical cervical cancer and an effective indicator for the molecular screening of cervical lesions [10].

The present study analysed the correlations between p16 expression and the clinicopathological features and prognosis of cervical cancer, to provide evidence-based medicine for the diagnosis and treatment of cervical cancer and important reference values for the development of clinical diagnostic and therapeutic decision-making methods for early-stage cervical cancer. The results provide a more targeted diagnosis and treatment of cervical pathology, improve the prognosis of cervical cancer, and provide valuable information for the diagnosis and treatment of cervical cancer in the clinic.

## 2 Material and methods

### 2.1 Inclusion and exclusion criteria

Literature inclusion criteria: (1) Study type: Clinical studies published in China and abroad on the differences in p16 expression in cervical cancer and its relationship with different clinicopathological features of cervical cancer in Chinese and English. (2) Study subjects: All cases had complete clinicopathological data, confirmed by pathological diagnosis, and had not received special treatment, such as surgery, radiotherapy or chemotherapy, before sampling. The control group included normal cervical tissues that were obtained during hysterectomy for other reasons (i.e., benign lesions). The included populations were not restricted by race, nationality, or age. (3) Case-control studies were included. (4) p16 protein was detected via immunohistochemistry. (5) The experimental data were complete and provided the number of cases and positive rate. (6) There were ≥20 patients in the experimental group and ≥15 patients in the control group. Literature exclusion criteria: (1) Reviews, abstracts, commentaries, and lectures. (2) Repeated publications by the same authors. (3) Studies with incomplete data or poor experimental design. (4) The detection method was not immunohistochemistry. (5) Animal experiments.

### 2.2 Bibliographic search

Computerised searches were performed for relevant literature in English databases (PubMed, Embase, Cochrane Library, and Web of Science) and Chinese databases (CNKI, Wanfang, CBM, and VIP databases). Case‒control studies published on the correlation between cervical cancer and the p16 protein were searched from January 01, 2010 to March 01, 2024.The literature retrieval personnel are LC and ZWZ. The following subject terms and free words were searched: cervical cancer subject terms+free words connected by OR, p16 subject terms+free words connected by OR, case-control studies connected by OR, and finally AND to connect the three parts for searching. The literature lists of eligible articles were filtered to locate other relevant articles. The search strategy for Chinese-language papers was similar to the English-language papers, with the publication language limited to English or Chinese, using the following search strategy. Step1: Cervical cancer subject words+free words are joined with OR (“Uterine Cervical Neoplasms” OR “Cervical Neoplasm, Uterine” OR “Cervical Neoplasms, Uterine” OR “Neoplasm, Uterine Cervical” OR “Neoplasms. Uterine Cervical” OR “Uterine Cervical Neoplasm”). Step 2: p16 subject words + free words are joined by OR (“Genes, p16” OR “Genes, p16INK4A” OR “p16INK4A Gene” OR “p16 Gene” OR “p16INK4”) Step 3: Case-control study. Step 4: The above three steps were searched by concatenating the terms with AND. No further ethical approval is required since the program does in review not require the recruitment of patients and the collection of personal information. This study has been granted the international PROSPERO website registration number: CRD42024546241.

### 2.3 Extraction of data and evaluation of the quality of literature

Two investigators independently extracted relevant data using a pre-designed form. Any disagreements were resolved via discussion. The following information was extracted from each study: serial number, name of the first author, year of publication, age range of the patients, FIGO stage, lymph node metastases of the patients, vascular infiltration, pathological grading, and clinical stage (Table 1). Study quality was assessed according to the Newcastle-Ottawa quality-assessment scale. Quality scores ranged from 0 to 9. The scale consists of three parts: the selection of research subjects (4 points), the comparability between the experimental group and the control group (2 points), and the measurement of outcome indicators (3 points). A total score of 0 - 5 points is regarded as a low - quality literature, while a score of 6 - 9 points indicates a high - quality literature. If the score is ≥ 6, it is considered a literature with high - quality. The work of extraction of data was operated by 2 independent researchers (LC and ZFD). Any disagreement was discussed and resolved with a third author (ZWZ). Extraction period: March to June 2024.

**Table 1.** Characteristics and results of the included studies.

| Study | Year | Country | Cervical cancer group (n) |  | Control group (n) |  | Method | Cervical cancer group |  |  |  |
| --- | --- | --- | --- | --- | --- | --- | --- | --- | --- | --- | --- |
|  |  |  | positive cases | Total | positive cases | Total |  | Age | Sample | Stage | Clinicopathological variables |
| Qiyang LI[21] | 2021 | China | 41 | 42 | 3 | 30 | IHC | 35-59 | SCC&ADC | I-III | D, DI, LN, PT, V |
| Xin ZHANG[22] | 2020 | China | 134 | 145 | 26 | 75 | IHC | 23-75 | ADC | I-II | D, DI, S1, LN, TS, V |
| Weiyang LI[23] | 2019 | China | 104 | 116 | 9 | 100 | IHC | 26-62 | SCC&ADC&AC | I-II | D, LN, S |
| Wenting LI[24] | 2019 | China | 48 | 50 | 12 | 50 | IHC | UN | SCC | I-II | D, LN, |
| Niuyan XUE[25] | 2019 | China | 55 | 100 | 14 | 100 | IHC | 31-69 | UN | I-IV | A, D, LN, |
| Ying LU[26] | 2018 | China | 24 | 40 | 1 | 40 | IHC | 34-65 | UN | I-IV | A, D, LN, |
| Yunxia LI[27] | 2017 | China | 92 | 100 | 3 | 50 | IHC | 23-66 | UN | I-II | D, DI, LN, S |
| Tingting TONG[28] | 2017 | China | 121 | 182 | 1 | 33 | IHC | 28-80 | SCC | I-IV | A, D, LN, |
| Ran YANG[29] | 2015 | China | 121 | 150 | 7 | 60 | IHC | 31-49 | SCC&ADC | I-IV | A, D, LN, PT, |
| Lina LIU[30] | 2015 | China | 38 | 40 | 0 | 15 | IHC | 41(average) | SCC | I-II | D |
| Diling PAN[31] | 2015 | China | 49 | 49 | 1 | 21 | IHC | 26-68 | SCC | I-II | LN, |
| Shrestha, S[32] | 2014 | China | 51 | 53 | 0 | 20 | IHC | 27-61 | SCC | I-III | LN, |
| Lei LEI[33] | 2014 | China | 23 | 52 | 4 | 26 | IHC | 32-85 | UN | I-IV | A, D, LN, |
| Yaping WANG[34] | 2013 | China | 22 | 30 | 0 | 15 | IHC | 40-69 | SCC | I-III | D, LN, V |
| Tiejun ZHOU[35] | 2013 | China | 26 | 36 | 4 | 15 | IHC | 26-63 | ADC | I-II | D, LN, V |
| Tao JIANG[36] | 2012 | China | 44 | 56 | 3 | 30 | IHC | 26-73 | SCC | I-II | A, D, LN, |
| Shuang LIU[37] | 2012 | China | 49 | 61 | 8 | 34 | IHC | 31-72 | ADC | I-IV | D, DI, S2, LN, TS |
| Weng, M[38] | 2012 | China | 38 | 62 | 1 | 20 | IHC | 24-78 | ADC | I-IV | D, S2, LN, PT, |
| Lili YAO[39] | 2011 | China | 21 | 23 | 0 | 20 | IHC | 22-55 | UN | I-II | A, D, DI, LN, TS |
| Yuyang ZHANG[40] | 2011 | China | 57 | 72 | 2 | 28 | IHC | 34-75 | SCC | I-IV | S2, LN, TS |
| Xiaoying XU[41] | 2011 | China | 55 | 62 | 1 | 22 | IHC | 25-70 | SCC&ADC | I-III | D, LN, PT, |
| Lu LIU[42] | 2010 | China | 35 | 38 | 5 | 20 | IHC | 30-70 | UN | I-IV | S2, LN, |
| Shanshan YANG[43] | 2010 | China | 48 | 52 | 2 | 28 | IHC | 31-67 | SCC&ADC | I-II | A, D, LN, PT, |

Markers: A, age( $\leq 40$ years VS $> 40$ years); D, histological differentiation (High + Medium VS Poorly); ADC, adenocarcinoma; AC, adenosquamous carcinoma; SCC, squamous cell carcinoma; DI, depth of tumor invasion ( $< 1/2$ VS $\geq 1/2$ ); IHC, immunohistochemistry; LN, lymph node metastasis(with VS without); S2( I-II VS III-IV stage); NOS, Newcastle-Ottawa quality assessment scale; PT, pathological type (squamous cell carcinoma VS adenocarcinoma); S, survival information; TS, tumor size( $< 4\text{cm}$ VS $\geq 4\text{cm}$ ) ; UN, unclear; V, vascular invasion (with VS without).

### 2.4 Statistical analysis

Review Man 5.4 software and STATA 16.0 software were used for systematic evaluation, and the results are expressed as ORs and 95% CIs as effect sizes. P<0.05 was considered a statistically significant difference. The heterogeneity of the extracted findings was tested using the I^2^ test. When P>0.1 and I^2^ ≤50%, there was no significant heterogeneity between the results of the studies, which were analysed using the fixed-effects model. I^2^ > 50% indicated heterogeneity between the results of the studies, which was analysed using the random-effects model. Review Man 5.4 software was used for statistics and data processing, and heterogeneity tests and sensitivity analyses were performed. STATA 16.0 software was selected for sensitivity analysis of the included studies, and Begg’s test was used to examine publication bias.

### 2.5 Selection of eligible studies

A total of 3209 articles were retrieved from the electronic search, 721 duplicates were removed, 43 reviews, reports, systematic reviews and Meta-analyses were removed, 16 animal experiments were removed, 2,396 studies that did not conform to the study after reading the abstract were removed, 6 cohort studies[11–16] were removed, 2 studies[17,18] with statistical errors and 2 studies[19,20] that did not give the complete outcome variable, leaving 23 studies[21–43] that were included in the study. Finally, twenty-three case control studies met the inclusion criteria. The details of the study selection process are presented in Fig 1. The characteristics and outcomes included in the study are presented in table 1. The reason of final exclusion of 10 articles is in table 2. The quality assessment table for the included research literature is presented in table 3.

**Fig. 1.**
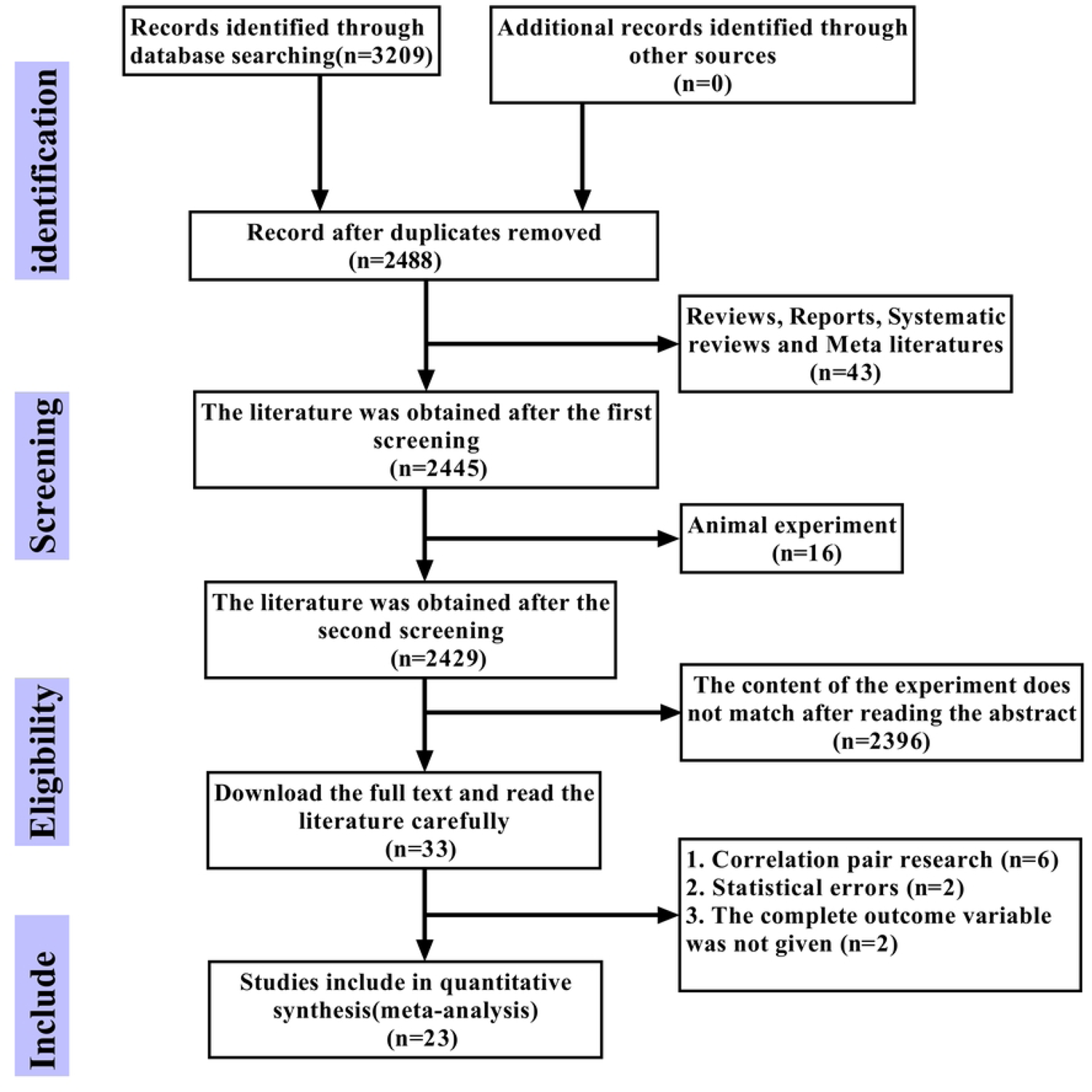
The study selection process

**Table 2.** The reason of final exclusion of 10 articles.

| Reference Number | Reasons for exclusion | Author | Year of Publication | Title of the Article |
| --- | --- | --- | --- | --- |
| 11 | Cohort study, without a control group. The included studies are all case-control studies. | Y.M. Chen<sup>[11]</sup> | 2010 | Expression and Clinical Significance of P16 and P15 Proteins in Cervical Cancer |
| 12 | Cohort study, without a control group. The included studies are all case-control studies. | W.H. Xie<sup>[12]</sup> | 2013 | Expression and Significance of p16 Gene in Cervical Intraepithelial Neoplasia and Squamous Cell Carcinoma Tissues |
| 13 | Cohort study, without a control group. The included studies are all case-control studies. | W.P. Wang<sup>[13]</sup> | 2013 | Expression and Significance of p16 and CXCR4 in Cervical Squamous Cell Carcinoma |
| 14 | Cohort study, without a control group. The included studies are all case-control studies. | C. Yue<sup>[14]</sup> | 2014 | Expression and prognosis significance of P16 in cervical cancer |
| 15 | Cohort study, without a control group. The included studies are all case-control studies. | J. Na<sup>[15]</sup> | 2019 | Association between galectin-1 and p16 protein expression and clinical characteristics of cervical cancer |
| 16 | Cohort study, without a control group. The included studies are all case-control studies. | Y.J. Zhang<sup>[16]</sup> | 2021 | Clinicopathological Characteristics of Cervical Cancer Patients and Analysis of P16, Stromal Cell-Derived Factor-1, Matrix Metalloproteinase-9, and Vascular Endothelial Growth Factor-C Expression |
| 17 | Statistical errors have occurred. | P. Deng<sup>[17]</sup> | 2021 | The Expression and Clinical Significance of p16, p53, and Ki67 in Cervical Cancer |
| 18 | Statistical errors have occurred. | J. Zhao<sup>[18]</sup> | 2019 | Analysis of the Expression and Pathological Features of p16, p53 and Ki - 67 Proteins in Cervical Cancer Tissues |
| 19 | The complete outcome variable was not given | Z. Chan-ping<sup>[19]</sup> | 2018 | **Expression and clinical significance of P16INK4A and KISS-1 in cervical cancer** |
| **20** | The complete outcome variable was not given. | W.U. Yi<sup>[20]</sup> | 2010 | Expression of Bmi-1 and p16 in cervical carcinoma carcinogenesis progression and its significance |

**Table 3.** Literature Quality Evaluation NOS Quality Evaluation Form.

| NO | Author and Year | Selection | Comparability | Exposure | Total NOS |
| --- | --- | --- | --- | --- | --- |
| 1 | Qiyang LI 2021 | ★★★ | ★★ | ★★ | 7 |
| 2 | Xin ZHANG 2020 | ★★★ | ★★★ | ★★ | 8 |
| 3 | Weiyang LI 2019 | ★★★ | ★★ | ★★ | 7 |
| 4 | Wenting LI 2019 | ★★★ | ★★ | ★★ | 7 |
| 5 | Niuyan XUE 2019 | ★★★ | ★★ | ★★ | 7 |
| 6 | Ying LU 2018 | ★★★ | ★★ | ★★ | 7 |
| 7 | Yunxia LI 2017 | ★★★ | ★★ | ★★ | 7 |
| 8 | Tingting TONG 2017 | ★★★ | ★★ | ★★ | 7 |
| 9 | Ran YANG 2015 | ★★★ | ★★ | ★★ | 7 |
| 10 | Lina LIU 2015 | ★★★ | ★★ | ★★ | 7 |
| 11 | Diling PAN 2015 | ★★★ | ★★ | ★★ | 7 |
| 12 | Shrestha, S. 2014 | ★★★ | ★★ | ★★ | 7 |
| 13 | Lei LEI 2014 | ★★★ | ★★ | ★★ | 7 |
| 14 | Yaping WANG 2013 | ★★★ | ★★ | ★★ | 7 |
| 15 | Tiejun ZHOU 2013 | ★★★ | ★★ | ★★ | 7 |
| 16 | Tao JIANG 2012 | ★★★ | ★★ | ★★ | 7 |
| 17 | Shuang LIU 2012 | ★★★ | ★★ | ★★ | 7 |
| 18 | Weng, M 2012 | ★★★ | ★★★ | ★★ | 8 |
| 19 | Lili YAO 2011 | ★★★ | ★★ | ★★ | 7 |
| 20 | Yuyang ZHANG 2011 | ★★★ | ★★ | ★★ | 7 |
| 21 | Xiaoying XU 2011 | ★★★ | ★★ | ★★ | 7 |
| 22 | Lu LIU 2010 | ★★★ | ★★ | ★★ | 7 |
| 23 | Shanshan YANG 2010 | ★★★ | ★★ | ★★ | 7 |

## 3. Results

### 3.1 p16 protein expression (cervical cancer patients VS normal controls) Meta-analysis

Analysis of the 23 included studies using ReviewMan 5.4 software showed OR = 48.25, I^2^ = 70% > 50%, P < 0.00001, with strong heterogeneity, and sensitivity analyses were needed to look for heterogeneity (Fig. 2A). Conducting a sensitivity analysis revealed significant heterogeneity among the studies conducted by LeiLEI, NiuyanXUE, TiejunZHOU, ShuangLIU, and XinZHANG (Fig. 2B), with I² = 70% > 50%, P < 0.00001. Consequently, after excluding these five studies, a subsequent heterogeneity assessment indicated that the remaining 18 studies exhibited no heterogeneity, with I² = 13%, P = 0.30 > 0.10. This suggests the absence of heterogeneity, allowing for the application of a fixed-effects model in the meta-analysis (Fig. 2C). In the cervical cancer cohort, the expression of p16 was found to be 74.32 times greater than that of the control group (OR = 74.32, 95% CI: 50.35-109.69, P < 0.00001), as detailed in table 4.

**Fig. 2.**
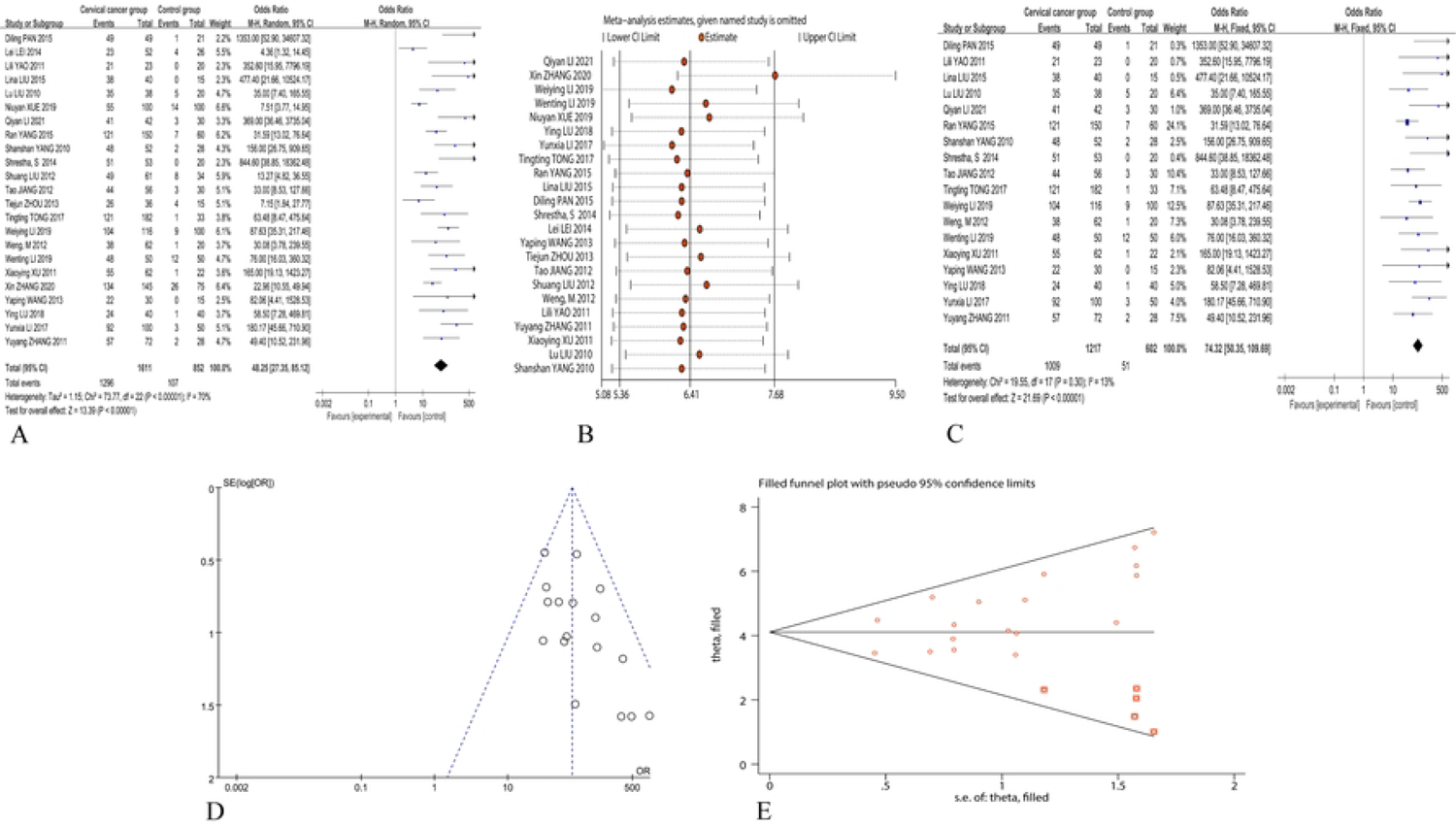
Meta-analysis forest plot of p16 expression in cervical cancer group compared to normal control group; forest plot of 23 studies (A); sensitivity analysis plot of 23 studies (B); forest plot of remaining 18 studies after excluding 5 studies with strong heterogeneity (C); funnel plot of remaining 18 studies (D); trim and fill plot of remaining 18 studies (E).

**Table 4.** Cervical cancer group vs. normal control group p16 expression positive.

| Sensitivity analysis | The number of study | Test for overall effect |  |  |  | Heterogeneity |  |  | Effect of the model | Publication bias ( Begg's test ) |
| --- | --- | --- | --- | --- | --- | --- | --- | --- | --- | --- |
|  |  | OR | 95% CI | Z | P | Chi<sup>2</sup> | P | I<sup>2</sup> (%) |  | P value |
| Including Lei LEI, Niuyan XUE, Tiejun ZHOU, Shuang LIU, Xin ZHANG study | 23 | 48.25 | 27.35-85.12 | 13.39 | < 0.01 | 73.77 | < 0.01 | 70 | Random | 0.003 |
| Excluding Lei LEI, Niuyan XUE, Tiejun ZHOU, Shuang LIU, Xin ZHANG study | 18 | 74.32 | 50.35-109.69 | 21.69 | < 0.01 | 19.55 | 0.30 | 13 | Fixed | 0.004 |

Funnel plots were generated using Review Manager 5.4.1 software to examine the presence of publication bias in this study. A symmetrical funnel plot indicates the absence of publication bias. The funnel plot for this study is presented in (Fig. 2D). Furthermore, a Begg’s test was conducted using STATA 16.0 software to validate the findings, yielding a P-value of 0.004, which is less than 0.05.

This suggests that the 18 selected studies in this research still exhibit a certain degree of publication bias. The existence of publication bias needs to be further addressed by using the trim-and-fill method on the asymmetric funnel plot above. The five red squares represent the effect sizes of the studies that need to be included in the future. By combining the funnel plot above, it can be seen that it is necessary to include five studies with results similar to those of the authors (Qiyan LI, Lina LIU, Diling PAN, Shrestha, S, Lili YAO) in order to ensure the symmetry of the funnel plot and eliminate publication bias (Fig. 2E). For specific analysis results (Table 4).

### 3.2 In cervical cancer (lymph node metastasis vs. lymph node non-metastasis), Meta-analysis of p16 expression positivity

Among the 23 studies, one study did not provide relevant data, resulting in the inclusion of data from 22 studies. A meta-analysis was conducted using Review Manager 5.4.1 software: I² = 68% > 50%, P < 0.00001, indicating the presence of moderate heterogeneity, necessitating a sensitivity analysis to investigate the sources of heterogeneity (Fig. 3A).

**Fig. 3.**
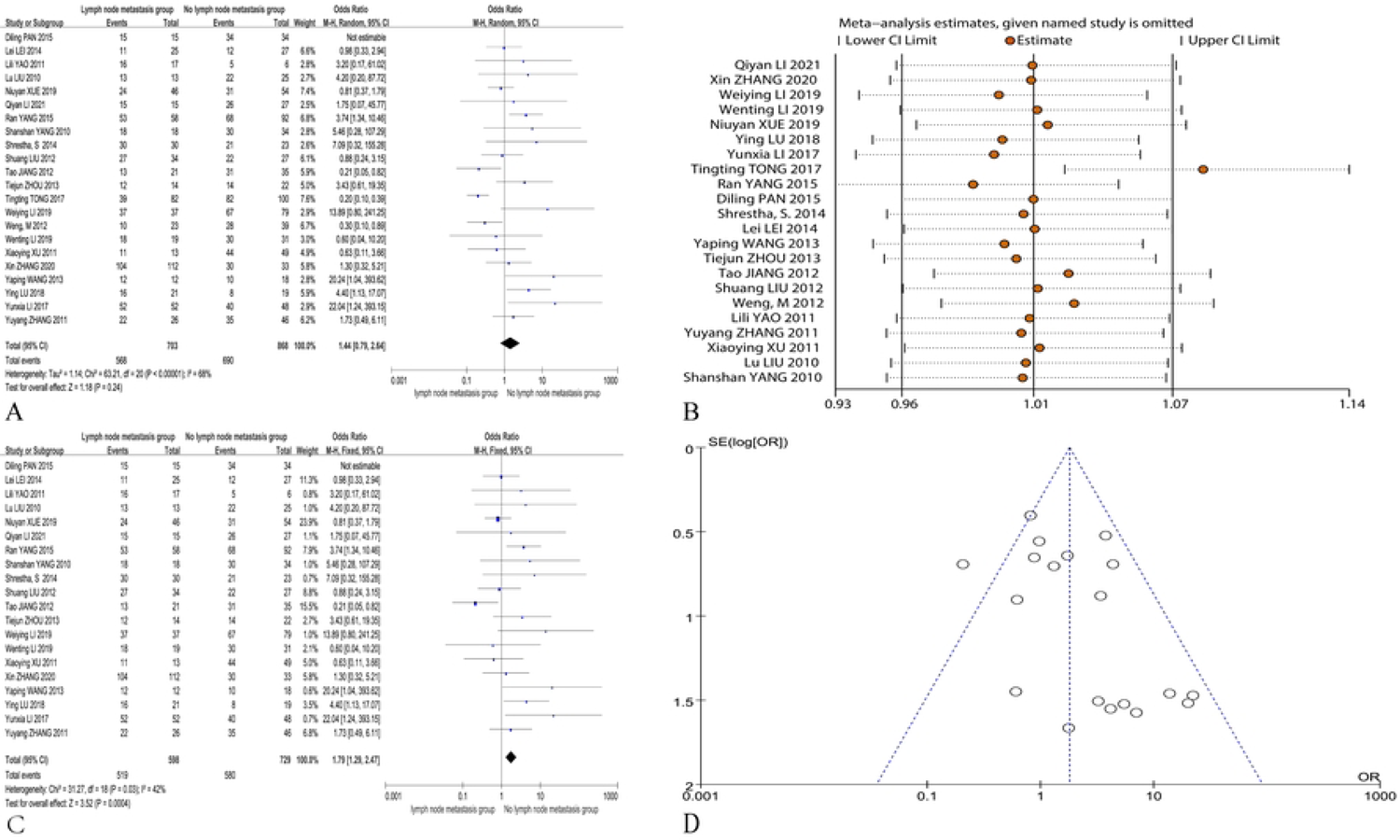
Meta-analysis of p16 expression positivity in cervical cancer (lymph node metastasis vs. non-lymph node metastasis): Forest plot of 22 studies (A); sensitivity analysis of 22 studies (B); forest plot of the remaining 20 studies after excluding 2 studies with high heterogeneity (C); funnel plot of the remaining 20 studies (D).

The analysis of the data from the 22 included studies revealed a significant heterogeneity, with I² = 68% > 50%, P < 0.00001 (Fig. 3B). Consequently, after excluding these two studies, a subsequent heterogeneity test was conducted, which indicated that the remaining 20 articles exhibited no heterogeneity, allowing for the application of a fixed-effects model for analysis (OR = 1.79, 95% CI: 1.29-2.47, P = 0.0004 < 0.05). This suggests that the expression of p16 in patients with lymph node metastasis in cervical cancer is 1.79 times that of patients in the non-metastatic lymph node group, with the difference being statistically significant. The forest plot and funnel plot were generated using Review Manager 5.4.1 software (Figs 3C and 3D).

The Begg’s test was conducted using STATA 16.0 software to verify the presence of publication bias in this study, with the results as follows: the funnel plot exhibited symmetry, and P = 0.234 > 0.05, indicating the absence of publication bias. Detailed analytical results are presented in table 5.

**Table 5.**
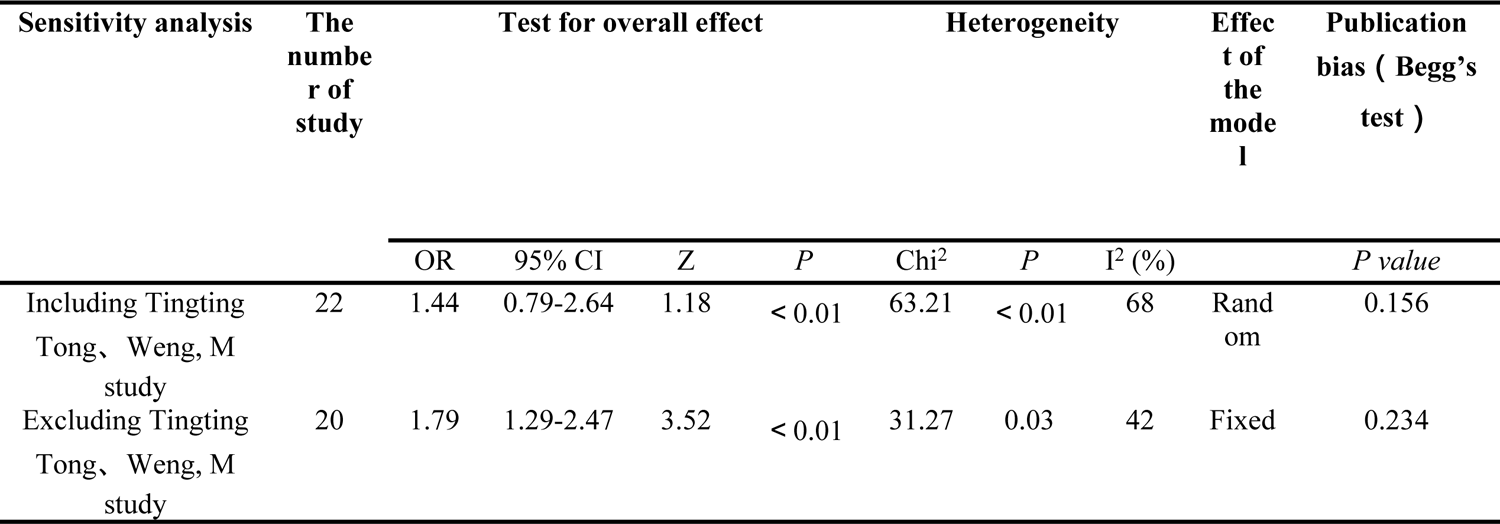
Meta-analysis and Sensitivity Analysis of p16 Expression Positivity in Cervical Cancer (Lymph Node Metastasis Group vs. Non-Metastasis Group)

### 3.3 Meta-analysis of p16 expression positivity in the cervical cancer group (high and middle differentiation group vs. low differentiation group)

A total of 19 studies provided data. The remaining 4 studies did not provide relevant data and were excluded. Meta-analysis was performed using Review Manager 5.4 software, which found I^2^ = 38% < 50%. A fixed effect model was used for analysis (OR=0.41, 95% CI: 0.30–0.56, P<0.00001), and the difference was statistically significant, which suggested that p16 expression in the high- and middle-level differentiation groups was 0.41 times greater than the low-level differentiation group among cervical cancer patients. The forest plots and funnel plots are as shown in Figs 4A and 4B.

**Fig. 4.**
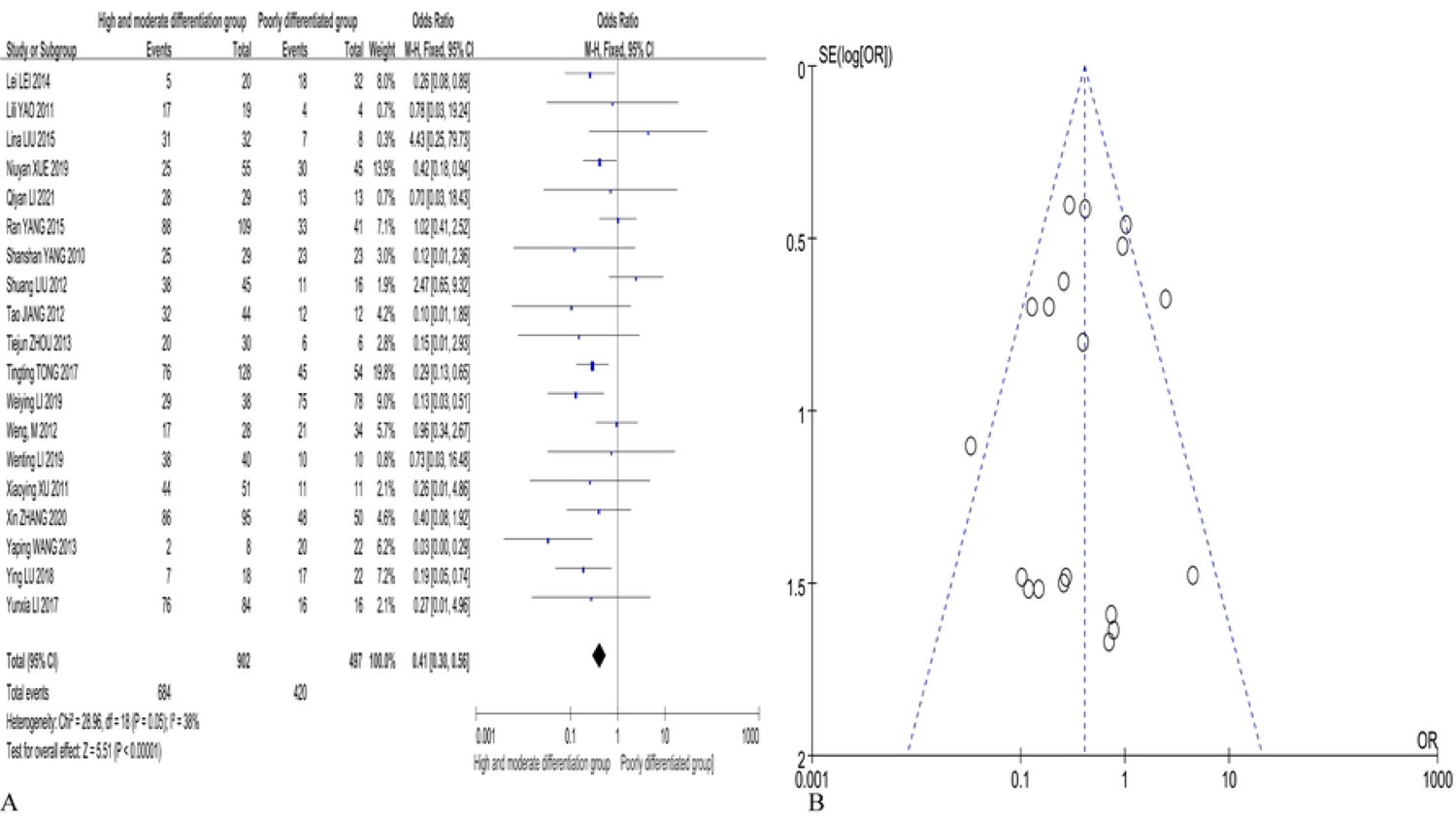
p16 expression in cervical cancer (high differentiated group VS low differentiated group) forest map(A);funnel map(B)

Bias test: STATA 16.0 software and Begg’s test were used to identify publication bias in this study, and the results revealed a symmetrical funnel plot (P=0.944 >0.05), which indicated no publication bias.

### 3.4 Meta-analysis of p16 expression positivity according to cervical cancer age (cervical cancer age ≤40 years vs. age >40 years)

Data from a total of 8 studies were included, and the remaining 15 studies did not provide relevant data or did not meet the inclusion criteria. Meta-analysis was performed using Review Manager 5.4 software, and I^2^ = 25% < 50%, P = 0.23 > 0.1, which suggested no heterogeneity. The fixed effects model was used for analysis. In the age≤40 years group vs. age>40 years group (OR=0.55, Z=3.00, P=0.003<0.05), the difference was statistically significant, which suggested that p16-positive expression in the cervical cancer age≤40 years group was 0.55 times greater than the age>40 years group. The forest plots and funnel plots are provided in Figs 5A and 5B.

**Fig. 5.**
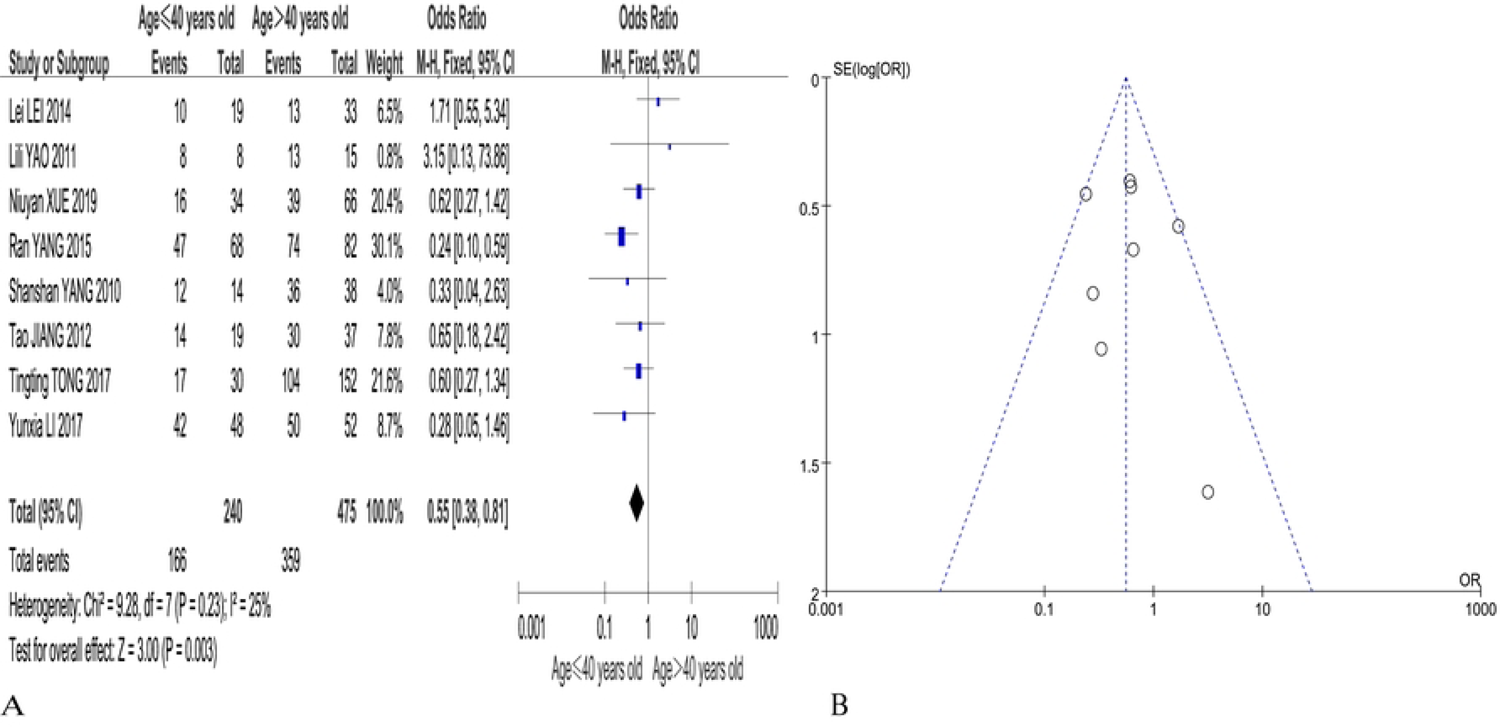
p16 expression in cervical cancer (Age≤40 years VS Age > 40 years) forest map (A);funnel map(B)

Begg’s test was performed using STATA 16.0 software to identify publication bias in this study, and the results revealed a symmetrical funnel plot (P=0.902 >0.05), which indicated no publication bias.

### 3.5 Meta-analysis of p16 expression positivity in cervical cancer according to FIGO stage (stage I-II vs. stage III-IV)

Data from 9 studies were included, and the remaining 14 studies did not provide relevant data. Meta-analysis was performed using Review Manager 5.4 software, and I^2^ = 36% < 50%, P = 0.20 > 0.1, which suggested no heterogeneity. A fixed-effects model was used for analysis, and the results revealed that the difference in tumour FIGO stage (stage I-II vs. stage III-IV) (OR = 0.37, 95% CI: 0.18-0.76, P = 0.0.007 < 0.05) was statistically significant, which suggested that the expression of p16 in the stage I-II group was 0.37 times greater than the stage III-IV group. The forest and funnel diagrams are provided in Figs 6A and 6B.

**Fig. 6.**
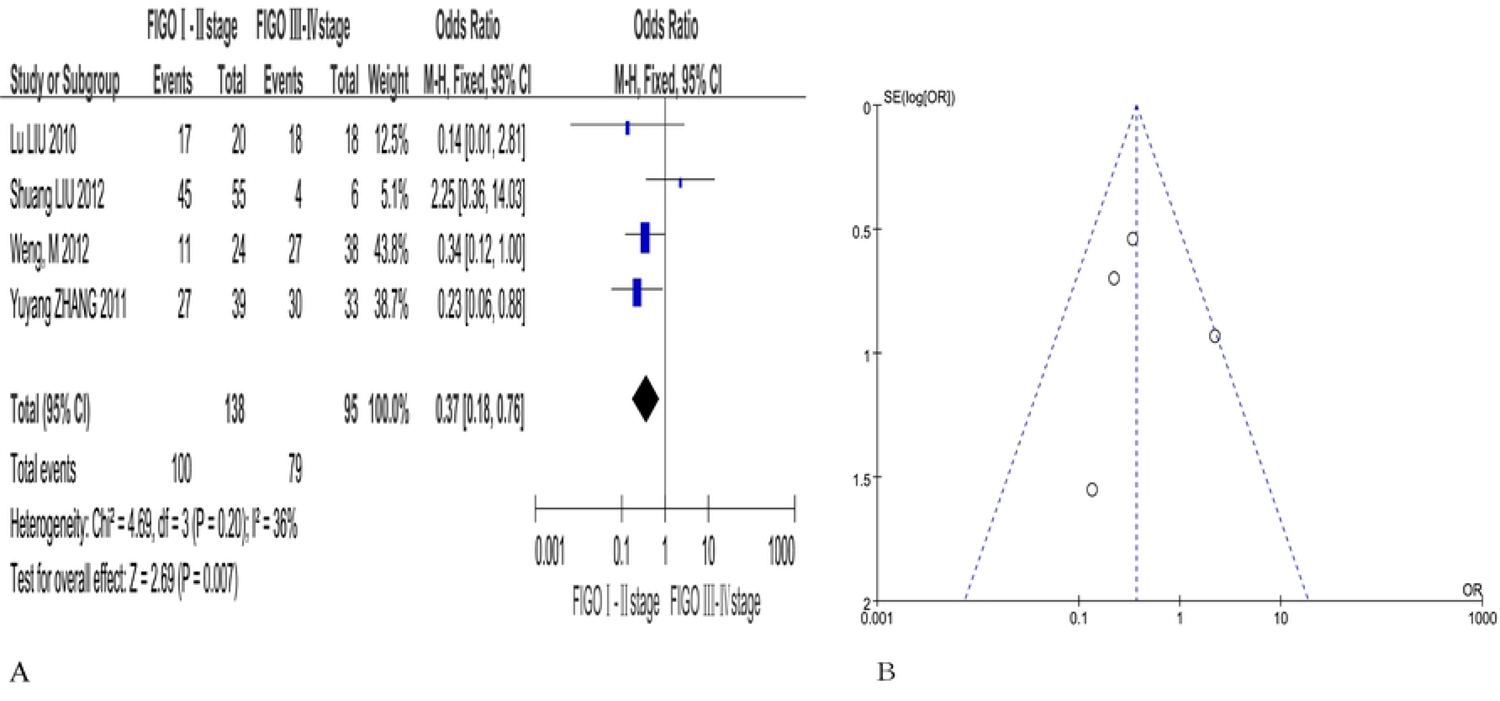
p16 expression in cervical cancer (Ⅰ-Ⅱstage VS Ⅲ-Ⅳ stage) forest map (A); funnel map(B)

Bias test: Begg’s test was performed using STATA 16.0 software to identify publication bias in this study, and the results revealed a symmetrical funnel plot (P=1.000>0.05), which indicated no publication bias.

### 3.6 Meta-analysis of p16 expression positivity according to the depth of infiltration of cervical cancer (<1/2 group vs. ≥1/2)

A total of five studies were included, while the remaining eighteen studies did not provide relevant data. Meta-analysis was performed using Review Manager 5.4 software, and I^2^ = 0% < 50%, P = 0.52 > 0.1, which suggested no heterogeneity. A fixed effects model was used for analysis, and the results revealed that the depth of cervical cancer infiltration (<1/2 group vs. ≥1/2) (OR=0.82, 95% CI: 0.40-1.71, P=0.60 > 0.05) was not statistically significant. The forest plots and funnel plots are shown in Figs7A and 7B.

**Fig. 7.**
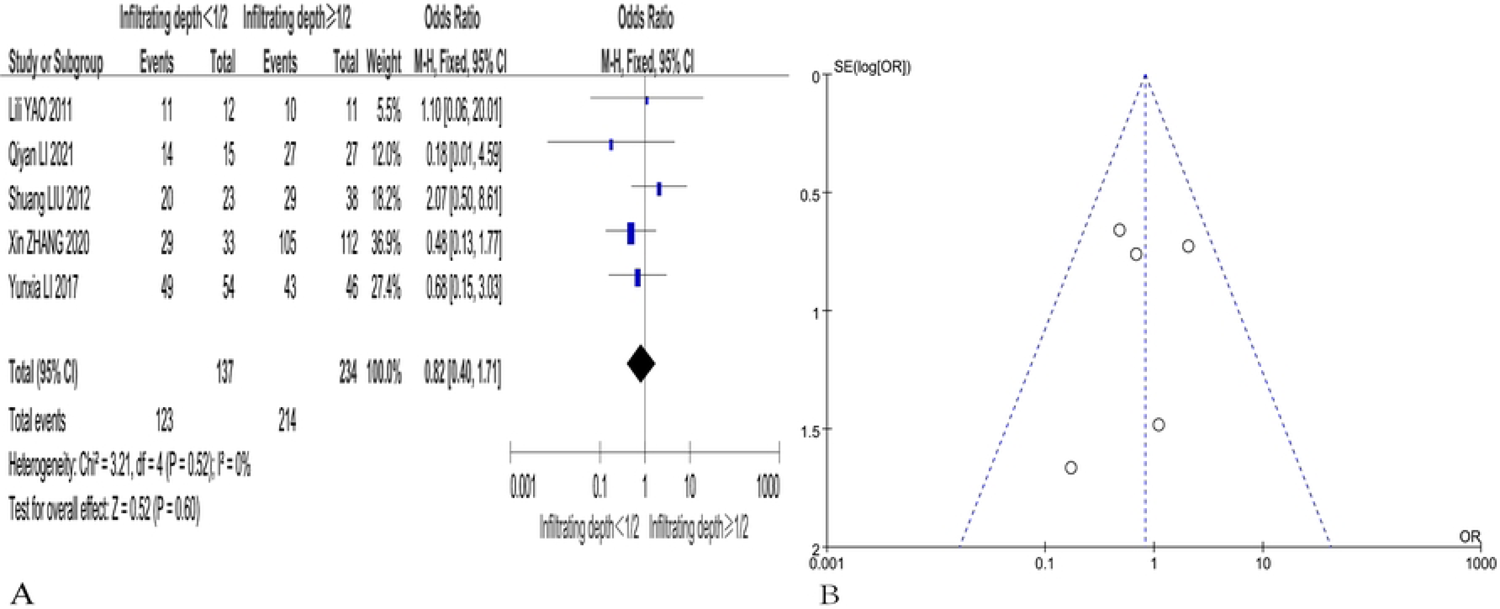
p16 expression in cervical cancer (<1/2 Depth of infiltration VS 1/2 Depth of infiltration) forest map(A);funnel map(B)

Begg’s test was performed using STATA 16.0 software to identify publication bias in this study, and the results revealed a symmetrical funnel plot (P=0.806 >0.05), which indicated no publication bias.

### 3.7 Meta-analysis of positive p16 expression in cervical cancer vascular infiltration (with vs. without)

Data from 4 studies were included, and the remaining 19 studies did not provide relevant data. Meta-analysis was performed using Review Manager 5.4 software, and I^2^ = 0% < 50%, P = 0.92 > 0.1, which suggested no heterogeneity. A fixed effects model was used for analysis, and the results revealed that for cervical cancer with vascular infiltration (with vs. without) (OR=3.53, 95% CI: 1.25-10.01, P = 0.02 <0.05), and the difference was statistically significant, which suggested that the p16 expression level in cervical cancer patients with vascular infiltration was 3.35 times greater than cervical cancer without vascular infiltration. The forest plots and funnel plots are shown in Figs 8A and 8B.

**Fig. 8.**
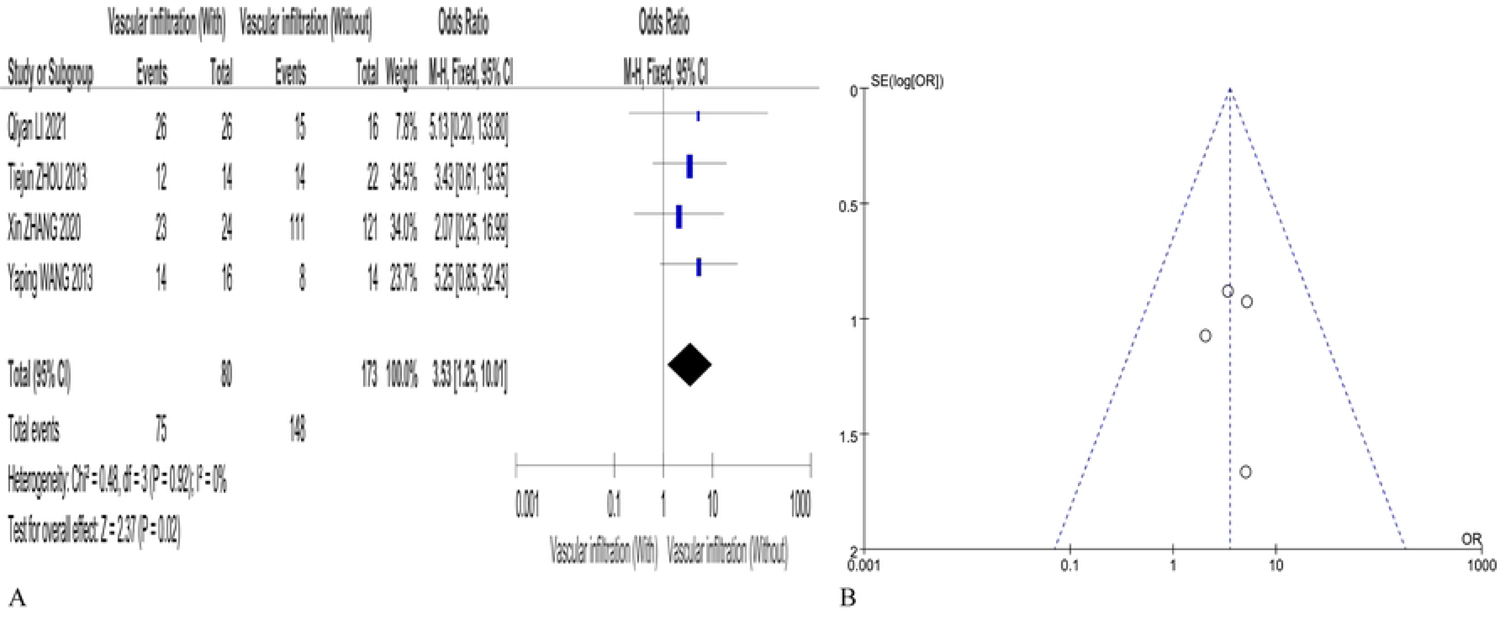
p16 expression in cervical cancer (With VS without vascular infiltration) forest map(A); funnel map(B)

Begg’s test was performed using STATA 16.0 software to identify publication bias in this study, and the results revealed a symmetrical funnel plot (P=1.0 >0.05), which indicated no publication bias.

### 3.8 Meta-analysis of p16 expression positivity according to cervical cancer pathological types (adenocarcinoma vs. squamous carcinoma)

Data from a total of 5 studies were included, and the remaining 18 studies did not provide relevant data. Meta-analysis was performed using Review Manager 5.4 software, and I^2^ = 0% <50%, P = 0.42 >0.1, which suggested no heterogeneity. A fixed effects model was used for analysis, and the results revealed that for the pathological type (adenocarcinoma vs. squamous carcinoma) (OR = 0.81, 95% CI: 0.40-1.65, P = 0.56 >0.05). The difference was not statistically significant, and the forest plots and funnel plots are provided in Figs 9A and 9B.

**Fig. 9.**
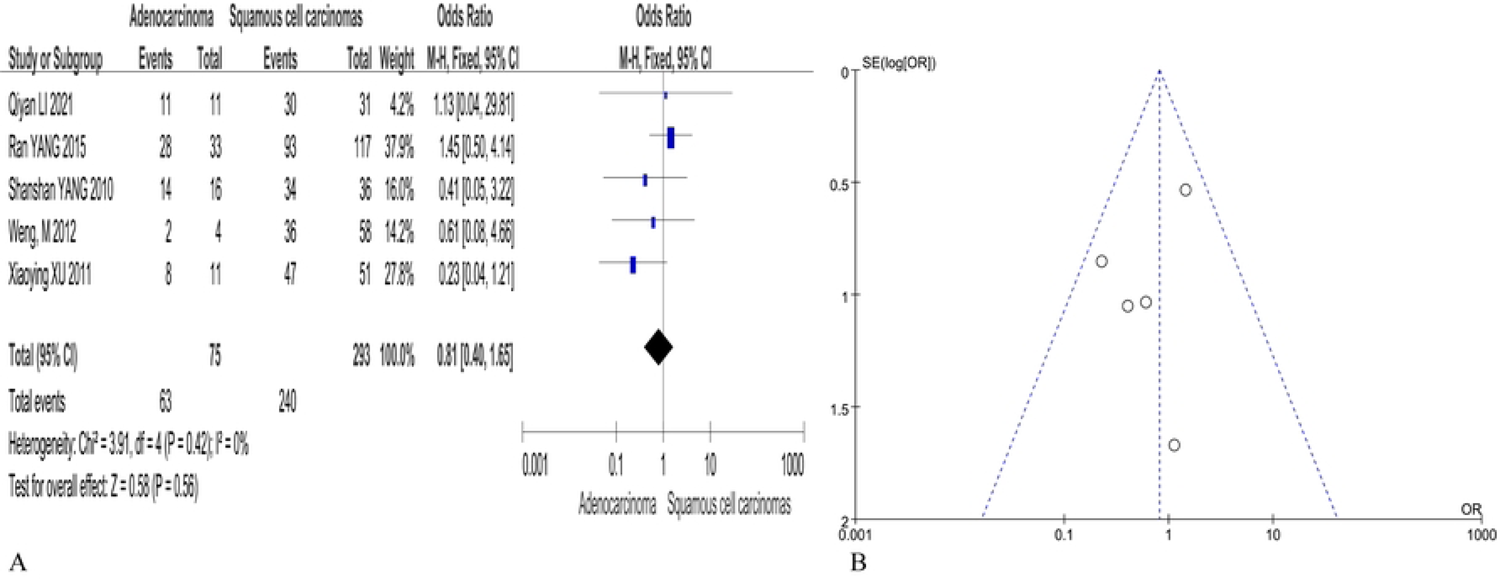
p16 expression in cervical cancer (Adenocarcinoma VS squamous cell carcinoma) forest map(A); funnel map(B)

Begg’s test was performed using STATA 16.0 software to identify publication bias in this study, and the results revealed a symmetrical funnel plot (P=1.0 >0.05), which indicated no publication bias.

### 3.9 Meta-analysis of p16 expression positivity according to cervical cancer tumour size (<4 cm vs. ≥4 cm)

Data from 4 studies were included, and the remaining 19 studies did not provide relevant data. Meta-analysis was performed using Review Manager 5.4 software, and I^2^ = 75% >50%, P = 0.007 <0.1, which suggested heterogeneity. A random effects model was used for analysis, and the results revealed that the difference in cervical cancer tumour size (<4 cm vs. ≥4 cm) (OR=0.80, 95% CI: 0.13-4.87, P=0.81>0.05) was not statistically significant. The forest plots and funnel plots are shown in Fig. 10A and Fig. 10B.

**Fig. 10.**
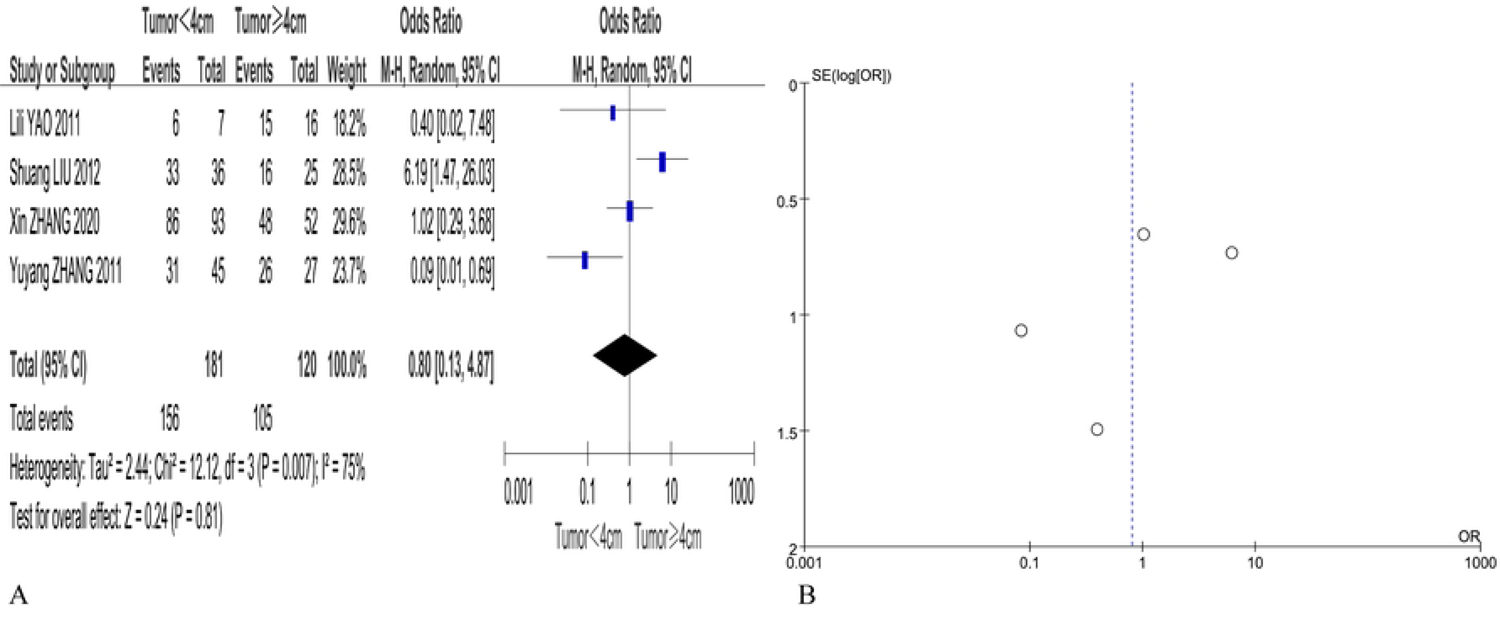
p16 expression in cervical cancer (Tumor size < 4cmVS≥4cm)) forest map(A); funnel map(B)

Begg’s test was performed using STATA 16.0 software to identify publication bias in this study, and the results revealed a symmetrical funnel plot (P=0.734 >0.05), which indicated no publication bias. The summary of the meta-analysis results regarding the relationship between p16 expression and clinical pathological characteristics is detailed in table 6.

**Table 6.** Meta-analysis evaluating the associations between p16 expression and clinicopathological variables.

| Clinicopathological variables of cervical cancer | The number of study | Cases of cervical cancer | Test for overall effect |  |  |  | Heterogeneity test |  |  | Effect of the model | Publication bias ( Begg's test ) |
| --- | --- | --- | --- | --- | --- | --- | --- | --- | --- | --- | --- |
|  |  |  | OR | 95% CI | Z | P value | Chi<sup>2</sup> | P value | I<sup>2</sup>(%) |  |  |
| The degree of differentiation ( High + Medium VS Poorly ) | 19 | 1399 | 0.41 | 0.30-0.56 | 5.51 | < 0.05 | 28.96 | 0.05 | 38 | Fixed | 0.944 |
| Age≤40 VS Age > 40 | 8 | 715 | 0.55 | 0.38-0.81 | 3.00 | < 0.05 | 9.28 | 0.23 | 25 | Fixed | 0.902 |
| FIGO staging ( I-II VSIII-IV ) | 4 | 233 | 0.37 | 0.18-0.76 | 2.69 | < 0.05 | 4.69 | 0.20 | 36 | Fixed | 1.000 |
| Depth of interstitial infiltration ( < 1/2VS≥1/2 ) | 5 | 371 | 0.82 | 0.40-1.71 | 0.52 | 0.60 | 3.21 | 0.52 | 0 | Fixed | 0.806 |
| Vascular invasion ( With VS Without ) | 4 | 253 | 3.53 | 1.25-10.01 | 2.37 | < 0.05 | 0.48 | 0.92 | 0 | Fixed | 1.000 |
| The pathologic types ( ADC VS SCC ) | 5 | 368 | 0.81 | 0.40-1.65 | 0.58 | 0.56 | 3.91 | 0.42 | 0 | Fixed | 1.000 |
| Tumor size < 4cmVS ≥4cm | 4 | 301 | 0.80 | 0.13-4.87 | 0.24 | 0.81 | 12.12 | 0.007 | 75 | Rando<br>m | 0.734 |

## 4 Discussion

The aetiology of cervical cancer is related to HPV infection in the cervix caused by multiple sexual partners and early sexual activity. Cervical cancer has an insidious onset and progressive disease development. The failure of patients to undergo timely diagnosis and effective treatment leads to further disease development and tumour cell migration into neighbouring tissues and organs throughout the body[44]. Neither cervical cancer screening nor pathological diagnosis can predict the future evolution of cervical lesions, but molecular markers are useful in the diagnosis of suspicious cases and in predicting the risk of recurrence or metastasis[45]. Accurate assessment of the severity and prognosis of cervical lesions can avoid overtreatment and reduce the psychological and economic burden on patients. Therefore, it is highly important to explore the expression of more objective molecular markers in cervical lesions to further elucidate the potential mechanisms of cervical cancer development and improve the diagnosis and treatment of cervical cancer.

p16 plays an inhibitory role in CDK4/CDK6 activity, prevents phosphorylation of the Rb protein, interferes with the dissociation of transcription factor (E2F), which prevents the transition from the G1 phase to the S phase and DNA synthesis to prolong the cell cycle. Therefore, organisms with abnormal p16 gene expression exhibit excessive cell division, and the absence of control over the cell cycle, which accelerates the rate of deterioration[46,47]. Several immunological studies showed that the p16 gene was not expressed in normal epithelium or benign lesions, but it was overexpressed in cervical dysplasia epithelial cells [48]. When a patient develops an HPV infection, p16 overexpression occurs due to the loss of negative feedback caused by the inactivation of pRb by the HPV oncoprotein E7[49]. In addition, Klaes R et al. [50] reported that the overexpression of p16 helped differentiate dysplastic cervical epithelial cells on cervical smears and histological sections. p16 is important in the detection and treatment of cervical cancer. Tsoumpou et al. [51] Systematically evaluated the correlation between published p16 immunostaining data and the degree of cytological or histological abnormality, and the results revealed that the proportion of p16 overexpression in cervical smears tended to increase with the severity of cytological abnormality. p16 diffuse staining was 82%, 68%, 38%, and 2% in CIN3, CIN2, CIN1 and normal cervical biopsies, respectively. p16/Ki67 cytological double staining increases the sensitivity of detecting high-grade precancerous lesions in cervical cancer, and it is a marker of transformed HPV infection[52]. The World Health Organization recommends the use of p16 immunostaining with HPV testing to differentiate between HPV-associated and HPV-independent cervical cancer.

In this study, 23 studies were meta-analyzed, and the results showed that p16 expression in cervical cancer was significantly higher than that in the normal control group, which was 74.32 times higher than that in the normal control group, and the difference was statistically significant, and the results of the studies were accordingly subjected to clipping and complementary experiments due to the publication bias of the results of the item studies, and the results of this study must be interpreted with caution. The results of this study indicate that the expression of p16 in cervical cancer patients is significantly associated with lymph node metastasis, tumor differentiation, age at diagnosis, FIGO staging of the tumor, and the presence of vascular invasion, with statistically significant differences observed. However, no significant correlation was found with cervical invasion depth, pathological type, or tumor size. The results of this Meta-analysis showed that high expression of p16 protein was a risk factor for lymph node metastasis, poor differentiation, old age of disease, late FIGO stage, and occurrence of vascular infiltration in cervical cancer. meta-analysis findings of K. Huang et al[53] found that p16 expression may predict a good prognosis for patients with cervical cancer, there are inconsistencies with the results of this experiment, which may imply that the expression of P16 protein needs to be controlled within a certain range. Exceeding a certain expression level may be associated with poor prognosis of cervical cancer, but large - scale, multi - center and well - matched cohort studies are required to confirm this finding.

There are certain limitations in this study. Among the 23 studies included, 22 were published in Chinese by Chinese researchers, and 1 was published in English by Chinese researchers on cervical cancer in China. No studies from other countries met the inclusion criteria. The article does not conduct direct research on cervical cancer patients but rather integrates studies from other scholars for a meta-analysis. Some conclusions within the text exhibit significant heterogeneity, prompting a comparative analysis before and after the exclusion of certain data. It is hypothesized that the substantial heterogeneity may be attributed to factors such as the wide variation in the age range of the included cases, the significant differences in the total number of cases, or the considerable discrepancies in the FIGO staging of the included cases. Furthermore, some of the research conclusions presented in the article exhibit publication bias, leading to the implementation of additional trim-and-fill analyses. The results indicate that future research must incorporate a greater number of studies to address this shortcoming, necessitating cautious interpretation of the findings. Therefore, it is essential for future studies to increase sample sizes and conduct multicenter research to supplement these deficiencies.

The World Health Organization has given a goal of 70% lifetime screening coverage to reach the goal of eliminating cervical cancer by 2030. There are several screening methods for cervical cancer, including visual inspection tests, cytology, human papillomavirus (HPV) DNA testing, and precision testing, which includes molecular and protein biomarkers, p16 immunostaining, HPV mRNA, and DNA methylation detection, to improve the specificity of screening[54].The clinicopathological features of cervical cancer and the degree of differentiation, age of disease, FIGO stage, and vascular infiltration are closely related to p16 expression. In the clinic, the infiltration, metastasis, and prognosis of cervical cancer can be evaluated according to p16 expression. The present findings provide valuable information concerning p16 as a potential target for targeted therapy in cervical cancer. HPV-positive cervical cancer exhibits cell cycle protein-dependent kinase (CDK) inhibition with increased p16 expression, which may be used as an alternative marker of HPV infection[55]. In future cervical cancer studies, investigators should be encouraged to report clinical outcomes, assess the overall response rate to cervical cancer treatment with new biomarkers, evaluate patient conditions, and identify more accurate prognostic factors.

High expression of P16 protein is a risk factor for lymph node metastasis, poor differentiation, advanced age at diagnosis, late FIGO staging, and vascular invasion in cervical cancer. However, P16 protein overexpression is not correlated with the depth of invasion, pathological type, or tumor size in cervical cancer. It is important to note that the prognostic assessment of cervical cancer is a multifactorial and comprehensive system; thus, relying solely on individual factors for prognostic evaluation is inappropriate. In clinical practice, for patients who are older, present with late FIGO staging, have larger tumor diameters, and exhibit positive lymph node metastasis and vascular invasion in postoperative pathology, a comprehensive assessment based on individual circumstances should be conducted. Necessary adjunctive therapies should be implemented with the aim of improving survival rates and enhancing prognosis for cervical cancer patients.

In conclusion, the p16 protein is closely related to the occurrence and development of cervical cancer and can serve as a reference indicator for the early warning and diagnosis of cervical cancer. The expression of p16 within a certain range may indicate a good prognosis for patients with cervical cancer, while its up - regulation may be associated with a poor prognosis of cervical cancer. In the future, it is necessary to expand the sample size for further in - depth and long - term research.

## Data Availability

All relevant data are within the manuscript and its Supporting Information files.

## Supporting information

S1 Table. PRISMA 2020 checklist. (DOCX)

S2 File. Original data (Excel)

S3 File. Review protocol (PDF)

S4 The 23 studies identified for inclusion in this research (DOCX)

S5 File. The final exclusion of 10 articles (DOCX)

S6 File. The table1-6 (DOCX)

## Author Contributions

Conceptualization: Le Chong, Zuowei Zou, Luhua Xia, Xinhua Wang, Zhanfei Dong, Yanping Zhao, Youxiang Hou

Data curation: Le Chong, Zuowei Zou

Formal analysis: Le Chong, Luhua Xia, Xinhua Wang Funding acquisition: Luhua Xia, Youxiang Hou

Investigation: Le Chong, Zuowei Zou, Zhanfei Dong, Yanping Zhao

Methodology: Le Chong, Zuowei Zou, Luhua Xia Project administration: Luhua Xia, Xinhua Wang Resources: Zhanfei Dong, Yanping Zhao, Youxiang Hou Software: Le Chong, Zuowei Zou, Luhua Xia Supervision:Xinhua Wang, Zhanfei Dong, Yanping Zhao Validation: Le Chong, Zuowei Zou, Luhua Xia Visualization: Le Chong, Zuowei Zou, Youxiang Hou Writing - original draft: Le Chong

Writing - review & editing: Le Chong, Zuowei Zou, Luhua Xia

